# POC-CCA3: Reducing batch-to-batch variation in the WHO-endorsed POC-CCA for *Schistosoma mansoni* and improving test interpretation

**DOI:** 10.1101/2025.06.04.25328804

**Authors:** Mahbod Entezami, Elías Kabbas-Piñango, Moses Arinaitwe, Theresia Abdoel, Pytsje T. Hoekstra, Sergi Alonso, Moses Adriko, René Paulussen, Govert J. van Dam, Poppy H. L. Lamberton, Joaquin M. Prada

## Abstract

**Background:** World Health Organization (WHO) schistosomiasis control programmes typically rely on Kato-Katz microscopy (KK) to detect *Schistosoma mansoni* eggs, although KK has limited sensitivity, especially in low-intensity infections. The WHO-endorsed point-of-care circulating cathodic antigen test (POC-CCA) is more sensitive but lacks the complete specificity of KK and has suffered significant batch-to-batch variability, compromising reliability. We evaluated an updated version, POC-CCA3, with the aim of addressing this batch-to-batch variation.

**Methods:** The POC-CCA3 employs recombinant monoclonal antibodies for Schistosoma CCA detection. School-aged children (n=870) in moderate (Tororo) and high endemicity (Mayuge) settings in Uganda were tested using KK, POC-CCA, and POC-CCA3 tests over three consecutive days. Data were analysed using descriptive statistics and Bayesian Latent Class Analysis, estimating diagnostic performance including sensitivity, specificity, receiver operating characteristic curves, and batch-specific variation and day-to-day variation.

**Results:** Model median prevalence estimates were 91.4% (Mayuge) and 31.2% (Tororo). A single POC-CCA3 test at G-score threshold 4 achieved estimated sensitivity and specificity, respectively, 86.3% and 92.9% in moderate and 88.1% and 95.5% in high endemicity settings. POC-CCA3 has a small increase in the probability of meeting the WHO TPP for monitoring and evaluation compared to the original POC-CCA. Overall diagnostic performance (AUC >94.2%) was higher compared to POC-CCA, with markedly lower batch-to-batch variation.

**Conclusions/Next steps:** The POC-CCA3 demonstrates superior performance and greater consistency compared to POC-CCA. A G-score threshold of 4 maximises performance towards WHO target product profile (TPP) requirements. However, neither test consistently meets WHO specificity requirements from a single sample, suggesting the need for continued refinement.

## 1 Introduction

Schistosomiasis, caused by trematodes of the Schistosoma genus, has the second greatest socio-economic impact of any parasitic disease after malaria [1]. It is endemic in 78 countries [2], with 90% of cases concentrated in sub-Saharan Africa [2, 3].

The World Health Organization (WHO) states that new diagnostics are needed for monitoring and evaluation (M&E) of ongoing national schistosomiasis control programmes and for use in interruption of transmission and surveillance scenarios [4]. WHO has released two target product profiles (TPPs) to guide the development and use of new or improved diagnostic tools for these two scenarios. More specifically, the TPP for M&E describes the need for diagnostic tools to capture when prevalence is above 10% in school-aged children, as communities above this threshold will require annual mass drug administration (MDA), and the diagnostic test would require a minimum of 60% sensitivity and 95% specificity for a sample of 100 people. For areas with low endemicity that are close to reaching interruption of transmission and post-elimination surveillance, the WHO TPP acknowledges that it may require a combination of two tests, with the first being highly sensitive (>99%), and the second test highly specific (>99%). If only one test is used for transmission interruption and surveillance purposes, 88% sensitivity and 99.5% specificity for a 3% prevalence threshold, with a sample of 100, is required [4]. Both TPPs have additional stringent requirements, and point-of-care tests with zero infrastructure needs, no or little sample processing, ease of use, and fast results are required for use in control and M&E settings. For interruption of transmission and post-elimination scenarios, it is acknowledged that other diagnostic tests with higher clinical sensitivity and specificity than point-of-care tests may be more likely to be able to meet the TPP requirements.

The primary species causing schistosomiasis in the African continent are *Schistosoma mansoni* and *Schistosoma haematobium* [5]. Adult Schistosoma worms infect humans, where they sexually reproduce and shed up to hundreds of eggs per day [6]. When excreted eggs come into contact with warm freshwater, they hatch to produce miracidia larvae, infecting snails where they reproduce asexually resulting in thousands of cercariae larvae, which are released back into the water. These cercariae burrow through the skin to infect humans before developing into feeding juvenile and adult worms to complete the life cycle. Adult worms lie in capillaries and feed off blood. As they do not have an anus, feeding waste is regurgitated [7], including circulating antigens that can be targeted for diagnosis [7, 8, 9]. A proportion of the eggs produced gets trapped in host tissues and stimulate immunological responses that are the main cause of morbidity [10]. The eggs that don’t get trapped are excreted in the stool (*S. mansoni*) or urine (*S. haematobium*) and cause transmission, but can also be used for infection detection. Adult worms are not accessible for direct diagnosis, and therefore all current diagnostic techniques provide indirect estimates of adult worm presence and numbers: eggs excreted in stool or urine, antigens regurgitated by feeding worms detected in urine or blood, DNA from different life-cycle stages, or host antibodies against the parasite.

WHO’s latest roadmap for neglected tropical diseases has targeted schistosomiasis for elimination as a public health problem (EPHP) in all endemic countries by 2030 [5]. MDA with the anthelmintic praziquantel has been the main control strategy for over 20 years. The main limitations of praziquantel are that the drug only acts on adult worms, meaning juvenile worms can still mature and reproduce, and it does not prevent reinfection, resulting in ongoing transmission soon after treatment [11, 12]. *S. mansoni* control programmes have historically relied on the WHO endorsed Kato-Katz thick smear microscopy (KK) to detect schistosome eggs in stool for infection diagnosis. KK has 100% specificity and requires no power source but has limited sensitivity, especially for low-intensity infections, is affected by daily variations in egg excretion [13] and non-homogeneous distribution within stool samples, requires well-trained technicians and microscopes, and is based on stool samples, which are commonly a less acceptable sample type than urine [14, 15].

The point-of-care circulating cathodic antigen test (POC-CCA), for the semi-quantitative detection of CCA [16] in urine, has been commercially available since 2008, and in 2017 was endorsed by the WHO as an alternative tool for the diagnosis of *S. mansoni* infections in endemic areas [18 - 19]. This test is used on urine samples, requires no transportation of samples, no additional equipment, minimal technical training, yields results in 20 minutes and is more sensitive than KK [14, 17], yet it does not have 100% specificity. Multiple studies have shown that the prevalence of *S. mansoni* infections measured by POC-CCA is substantially higher than KK-based prevalence [18, 19, 20, 21], with latent class analysis supporting the higher prevalence estimates and lower drug efficacy results, underscoring the limitations of KK [16, 22].

Endemicity levels and MDA frequency are categorised by WHO guidelines, based primarily on infection prevalences in school-aged children as measured by one KK [4]. Although the translation of threshold values between KK and POC-CCA for programme decision support was recommended by the Strategic and Technical Advisory Group for Neglected Tropical Diseases (STAG-NTD) in 2015 [23, 24], KK is still the primary tool in use today in part, due to the ability to compare new KK data with historical data, capability to score infection intensities, 100% specificity, highly trained personnel used to performing this test. However, POC-CCA was historically only scored as negative, ‘trace’, or positive, with a score of +, ++ or +++ for increasing intensity, but these scores, especially ‘trace’ are likely affected by inter-reader variation and lighting levels. The G-score system provides improved semi-quantitative measurements, using a score of 1-10, and these are aligned to printed cassettes, to reduce inter-reader variability [25]. However, the difficulties in interpreting some of the POC-CCA test results, in particular the ‘trace’ readings remain [26]. Furthermore, limitations have been compounded with issues in production reproducibility and quality, resulting in substantial batch-to-batch variations [27, 28]. The first POC-CCA test was performed with a chase buffer added after the urine sample, the second version incorporated this buffer straight into the test strip as a dry buffer to improve the field applicability of the test, however, this appeared also to lead to more batch-to-batch variation [27, 28]. Calibrator samples containing known CCA concentrations, and G-scores could be used to assess the consistency of different production batches and ensure accuracy in results. By comparing, for each calibrator, the G-score per batch with the baseline (predicted) G-score provided for the calibrators, any deviations or variations between batches can be identified. The calibrators facilitate the detection of skewed outputs and enable the standardisation of readings, accounting for batch-specific variations and ensuring the reliability and uniformity of the test results in the laboratory and the field.

The ideal scenario of using only one POC-CCA test per person does not enable day-to-day (the difference in G-score when sampling consecutive days) or within-sample variation (the difference in G-scores when testing the same sample) to be measured or controlled for and often only one batch of tests is used [16, 29, 30]. An improved POC-CCA that could reduce batch-to-batch variation is needed, and/or calibrators that could be used to control any remaining variation in the field. These calibrators could be used in combination with the G-scores to improve semi-quantitative infection intensity scoring as well as to reduce inter-reader bias, especially near the positive/negative threshold point [25].

Our overarching objective was to compare a new POC-CCA (called POC-CCA3) in comparison the existing POC-CCA in addressing the mention problems with batch-to-batch variation, in accordance with the M&E TPP guidelines. To assess this we focused on four specific objectives: 1) quantify batch-specific variations for the POC-CCA and POC-CCA3 test and the need for a set of calibrator samples to be included in the POC-CCA3 test kit to help reduce the impact of any potential batch-to-batch variation; 2) quantify the sensitivity and specificity of the POC-CCA3 in moderate and high *S. mansoni* endemic settings, compared to the existing POC-CCA and KK using latent class analysis; 3) compare different thresholds for both POC-CCA3 and POC-CCA to determine when they meet the TPP criteria, and to provide recommendations on appropriate G-score thresholds using POC-CCA3 to achieve the WHO TPP for monitoring and evaluation; 4) identify the POC-CCA3 prevalence in a population to align with current WHO guidance of 10% prevalence using single slide KK.

## 2. Methods

### 2.1 Laboratory evaluation

#### 2.1.1 POC-CCA3 production and quality control

To reduce the batch-to-batch variation seen in POC-CCA, we developed an improved version called POC-CCA3 (Mondial Diagnostics, The Netherlands, with support from the Leiden University Medical Center), using recombinant anti-CCA antibodies instead of the monoclonal anti-CCA antibodies used in the currently marketed POC-CCA version from Rapid Medical Diagnostics, South-Africa [31]. For the POC-CCA3, a recombinant version of the specific monoclonal antibody (human IgG) against Schistosoma CCA has been used as a capture, and detection antibody. The capture antibody and anti-human IgG antibodies were immobilised on the nitrocellulose membrane of the test line and control line, respectively, using a Biodot dispensing system. The detection reagent was prepared by conjugation of the specific monoclonal antibody to 40 nm colloidal gold particles, which was then applied to the conjugate pad. Following assembly of the nitrocellulose membrane with the sample, conjugate and absorbent pads on a backing card the master card was cut into 3.9 mm-wide strips, which were placed in plastic cassettes and individually sealed in an aluminium pouch containing a desiccant. A set of 3 lyophilized calibrators was provided for use with the POC-CCA3 kits, to assess if the G-scores of the calibrators can be used to adjust for inter-batch variations inter-reader bias. In total, three batches of 1,600 POC-CCA tests each were purchased from ICT International (Cape Town, Republic of South Africa). Three batches of 1,850 POC-CCA3 tests each were manufactured and provided by Mondial Diagnostics (Amsterdam, the Netherlands) (with 250 of these used with the calibrators). After receiving the batches, quality control of the three POC-CCA batches (Batch numbers: 220701075, 220902098 and 221117133) and the three POC-CCA3 batches (Batch numbers: SCHR2304-1, SCHR2304-2 and SCHR2304-3) was performed in the laboratory. For this quality control process, three replicates of CCA-negative urine samples spiked with 12 known concentrations of adult-worm-antigen trichloroacetic acid-soluble fraction AWA-TCA, containing approximately 3% of CCA [32, 33]. Concentrations were used. Final CCA concentrations in these AWA-TCA spike urines ranged from 0 to 240 ng/mL (0, 0.3, 1, 1.5, 2.1, 2.4, 2.7, 3.3, 3.9, 4.5, 24 and 240 ng/mL). Each of the POC-CCA and POC-CCA3 batches was tested in triplicate with all the spiked urine samples, according to the manufacturer’s instructions. The cassettes were read 20 minutes after adding the urine, using the G-scores (G1-G10) [25]. For evaluation, we considered G-scores of 2, 3, and 4 as thresholds for positive diagnosis.

#### 2.1.2 Modelling dose-response relationship

The dose-response relationship between G-score and CCA concentration for AWA spiked sample i, and each batch of POC-CCA and POC-CCA3, b, was assumed to follow a logistic curve, formulated by:

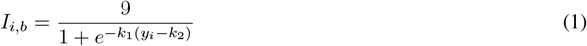

where *y*_*i*_ represents the CCA concentration of the sample *i*, the parameter *k*_1_ controls the steepness of the logistic curve, while *k*_2_ represents the midpoint concentration. *I*, here, is the colour intensity of the G-score band. Non-informative priors were assigned to *k*_1_ and *k*_2_. Each batch, *b*, was modelled independently to capture the potential variability in assay conditions across batches. The recorded G-score was then drawn from a normal distribution with a mean of *I*_*i,b*_ and a variance of *τ*_*b*_, representing the variance of batch *b*, drawn a non-informative prior with a gamma distribution as before. This is to simulate the variation seen between samples with the same concentration, either due to batch variation or reader bias.

### 2.2 Field evaluation

#### 2.2.1 Data collection

The data used in this study were collected from Mayuge District, Uganda, a high *S. mansoni* endemicity setting, and Tororo District, Uganda, a moderate *S. mansoni* endemicity setting during July 2023 (Mayuge) and September 2023 (Tororo). Eight schools in Mayuge and eight communities around schools in Tororo were selected and visited for sample collection. A total of 448 and 422 children were recruited in Mayuge and Tororo, respectively. Ages ranged between 6 and 13 years old in the schools surveyed in Mayuge, and between 6 and 14 in the schools in Tororo. One stool and one urine sample were collected per child per day for three consecutive days. Duplicate KK were prepared from each stool sample, and eggs were counted on microscopes at 10x magnification, resulting in six KK readings per child. Mean KK counts were multiplied by 24 to estimate eggs per gram of stool. On each urine sample from the three different days, both POC-CCA and POC-CCA3 were performed. Additionally, on the day-1 urine sample, two more replicates of the POC-CCA and POC-CCA3 tests were performed. Both were performed according to the manufacturer’s instructions. In short, each urine sample was stirred, and the Pasteur pipettes provided in the POC-CCA kits were used to deliver two drops (approximately 100 *µ*L) to the sample well of the test cassette. The POC-CCA3 kits included 100 *µ*L exact volume transfer pipettes to introduce the urine sample into the test cassette (used as per the instructions in the IFU) to provide a more accurate control of the sample volume. Intensities of the test lines were scored after 20 minutes, using G-scores. There were two technicians scoring each test, and a consensus on the G-score was reached and recorded. As an additional quality control, from each box of 25 POC-CCA3 tests, three tests were used with three calibrators provided by the manufacturer (1.8, 2.4 and 3 ng/*µ*L CCA).

Individuals were randomly assigned to one of three groups (A, B, or C), with each group corresponding to a specific allocation of test batches (Supplemental Material 1). On the first sampling day, all urine samples were tested using all three batches of both the POC-CCA and POC-CCA3 diagnostics. For Days 2 and 3, each individual was tested using only one batch per test, determined by their group allocation. Due to logistical constraints and occasional unavailability of certain batches, some individuals were tested using only two batches instead of three. This was accounted for in the model by incorporating the exact batch used for each sample.

After the final stool and urine sample had been collected, study participants were fed bread and juice before treatment (40 mg/kg praziquantel), irrespective of infection diagnosis result and as part of the national MDA programme.

To assess the agreement between the POC-CCA, POC-CCA3 and KK, two Spearman’s rank correlation analyses were performed. Firstly, the correlation between POC-CCA and POC-CCA3 readings G-scores was evaluated. This comparison aimed to establish the consistency between the assays by determining the strength of the relationship between their scores. Secondly, the relationship between the KK-derived egg counts and the POC-CCA3 G-scores was also investigated. For this purpose, average daily KK values were correlated with average daily POC-CCA3 G-scores.

#### 2.2.2 Latent class model analysis

A latent class model of *S. mansoni* infection in humans was developed from previous frameworks [11, 16, 26, 34] to estimate infection and diagnosis considering KK, POC-CCA, and POC-CCA3 as imperfect estimators of individual infection intensity. The true infection intensity was assumed to be a gamma distribution across the infected population. This framework enables the distinction between infected and uninfected individuals and the estimation of the infection status based on a combination of all diagnostic results. We can define individual infection status as:

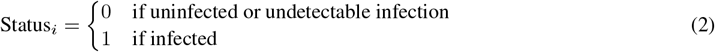

With *Status* = 0 meaning that the individual was uninfected or with an undetectable infection, and Status = 1 meaning that the individual was infected, for each individual *i*. Population prevalence is estimated by the proportion of individuals with *Status* = 1. Each individual had an intensity of infection, *λ*_*i*_, drawn from a gamma distribution of intensity across the population with a shape, *s*, and rate, *r*, with different values for Mayuge and Tororo:

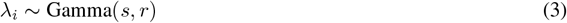

To account for the measurement errors possible in KK, a negative binomial distribution was used [34, 35], with an over-dispersion parameter *ω*. Individuals uninfected with schistosomiasis should express a KK count of zero (as the diagnostic is assumed to have 100% specificity). KK diagnosis for individual *i*, can thus be calculated as:

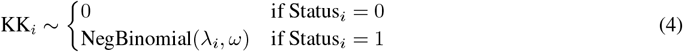

The POC-CCA test’s strength of colour is a quantitative ourcome from which the reader decifers a G-score, which is a semi-quantitative outcome, with possible scores between 1 - 10 as observed by the reader [25]. We assume the G-score to be drawn from the strength of colour, *I*, using a normal (Gaussian) distribution with a precision of 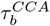 and 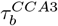 for each batch *b*, to account for inter-batch variability and the bias each time the technician matches the colour to a band of a G-score. The POC-CCA strength of colour, 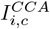, for each individual, *i*, and repeat sample, *c*, was determined by the individual’s intensity of infection, *λ*_*i*_, with a logistic function with model parameters 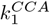 and 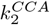 estimated:

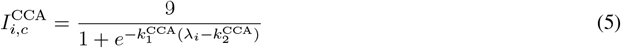

Similarly, the POC-CCA3 G-score was drawn from a normal distribution, with a precision of 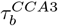. A similar logistic function is used to calculate the POC-CCA3 strength of colour, 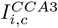, with the intensity of infection, *λ*_*i*_, and model parameters 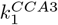 and 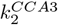 being estimated:

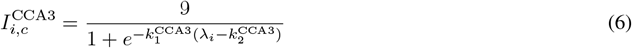

The model was run in a Bayesian framework package in R [36] using a Gibbs sampling algorithm (packages “rjags” [37] and “run-jags” [38]). A burn-in period of 1000 iterations was run with 5000 samples and two independent chains. Convergence was assessed by visual examination of the trace plots and the Gelman-Rubin statistic. The run includes all individuals, with up to 2 KK samples and one POC-CCA and POC-CCA3 value each for every one of the three days of sampling, along with 2 POC-CCA and POC-CCA3 replicates for the first day. We assessed the goodness-of-fit of the model by comparing draws from the posterior distribution to the data, see Additional file 1. All code generated is available in a GitHub repository (https://github.com/MabEntez/Schisto-CCA3-LCA).

The POC-CCA and POC-CCA3 portions of the model were initially fitted using results from the laboratory evaluation based on spiked samples. This enabled the comparison of batch-specific variation of both diagnostics. 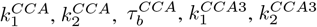, and 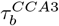 prior distributions were informed by previous models [26, 34]. After the model fitting, we obtained a posterior distribution for each of the fitted parameters (shapes and rates of the gamma distribution, 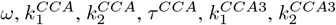, and 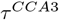). The posterior distributions of 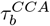 and 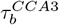, representing the batch-specific precisions for the POC-CCA and POC-CCA3 diagnostics, were used to assess the variability across batches under field conditions.

#### 2.2.3 Receiver Operating Characteristic (ROC) curves and Area Under Curve (AUC)

POC-CCA and POC-CCA3 generate G-scores based on the amount of CCA in the urine, reportedly linked to an individual’s infection intensity [39, 40, 41, 42]. Depending on the selected G-score threshold, these results are interpreted as either positive or negative for schistosomiasis. Different thresholds can yield varying sensitivity and specificity, particularly depending on the endemic setting. To quantify the sensitivity and specificity of each diagnostic test, we reconstructed the logistic functions and established the relationship between true infection intensity and expected G-scores for each diagnostic and setting. For each posterior, we simulated 1,000 individuals, predicting their infection status, infection intensity, and G-scores for each diagnostic. Sensitivity was calculated as the proportion of infected individuals with G-scores equal to or more than a threshold, while specificity was the proportion of uninfected individuals with G-scores below the threshold. A random batch of each diagnostic was assumed for each individual sample to account for variability between batches and to approximate the expected sensitivity and specificity profile in future applications. This approach aimed to reflect the average diagnostic performance across multiple batches, providing a more representative estimate of how these tests would perform in practice. We assessed the impact of three different G-score thresholds (2, 3, 4) over 1, 2, or 3 days of sampling. These sensitivity and specificity values were then used to create ROC curves and calculate AUC values. Finally, we determined the proportion of simulations that met the minimum (60% sensitivity, 95% specificity) and optimum (75% sensitivity, 96.5% specificity) WHO TPP for M&E requirements, considering the number of sampling days, endemicity setting, and G-score threshold.

Furthermore, we aimed to compare the prevalence estimated by POC-CCA and POC-CCA3 with a 10% KK prevalence using a single slide. To do so, we employed the model posteriors corresponding to the 100 lowest prevalence estimates (noting that in Tororo, a single slide KK yielded a prevalence of 14%; Table 1). We then simulated 10,000 populations by randomly sampling from these posteriors and calculating KK, POC-CCA, and POC-CCA3 prevalence estimates, retaining the 1,000 simulations that most closely approximated a 10% KK prevalence.

**Table 1:** Summary of descriptive results for Schistosoma mansoni prevalence, based on the average result per child across all collected samples: six samples for Kato-Katz (KK), five for the point-of-care circulating cathodic antigen test (POC-CCA), and five for the improved version of the same test with reduced batch-to-batch variation (POC-CCA3).

| Diagnostic | Metric | Mayuge (n=448) | Tororo (n=422) |
| --- | --- | --- | --- |
| Model | Model predicted prevalence (%) | 91 | 31 |
| KK | Prevalence using all samples (%) | 72 | 15 |
|  | Prevalence using the first sample (%) | 54 | 12 |
|  | Average intensity (eggs per gram) | 8.8 | 0.5 |
| POC-CCA with varying G-score thresholds | Prevalence average of all samples (%) | G2: 96.1, G3: 89.1, G4: 83.9 | G2: 60.1, G3: 28.7, G4: 19.1 |
|  | Average intensity using all samples (G-score) | 7.7 | 2.7 |
|  | Prevalence using the first sample (%) | G2: 99.3, G3: 93.2, G4: 90.3 | G2: 26.7, G3: 21.2, G4: 16.5 |
|  | Average intensity using the first sample (G-score) | 6.7 | 2 |
| POC-CCA3 with varying G-score thresholds | Prevalence average of all samples (%) | G2: 89.4, G3: 82.9, G4: 77.9 | G2: 38.3, G3: 28.6, G4: 25.7 |
|  | Average intensity using all samples (G-score) | 7 | 2.9 |
|  | Prevalence using the first sample (%) | G2: 88.7, G3: 80.0, G4: 83.9 | G2: 36.0, G3: 32.2, G4: 26.5 |
|  | Average intensity using the first sample (G-score) | 6.9 | 3 |

## 3 Results

### 3.1 Laboratory evaluation: quality control

Across the three batches, the POC-CCA3 exhibited an earlier rise in G-score at very low CCA concentrations compared with the POC-CCA (Figure 1). In addition, the POC-CCA3 data showed a narrower range of lines of best fit, indicating greater consistency across replicates, whereas the POC-CCA data displayed a wider spread. Despite these differences at the lower end of detection, both assays appeared to plateau, on average, in a comparable manner at higher concentrations.

**Figure 1:**
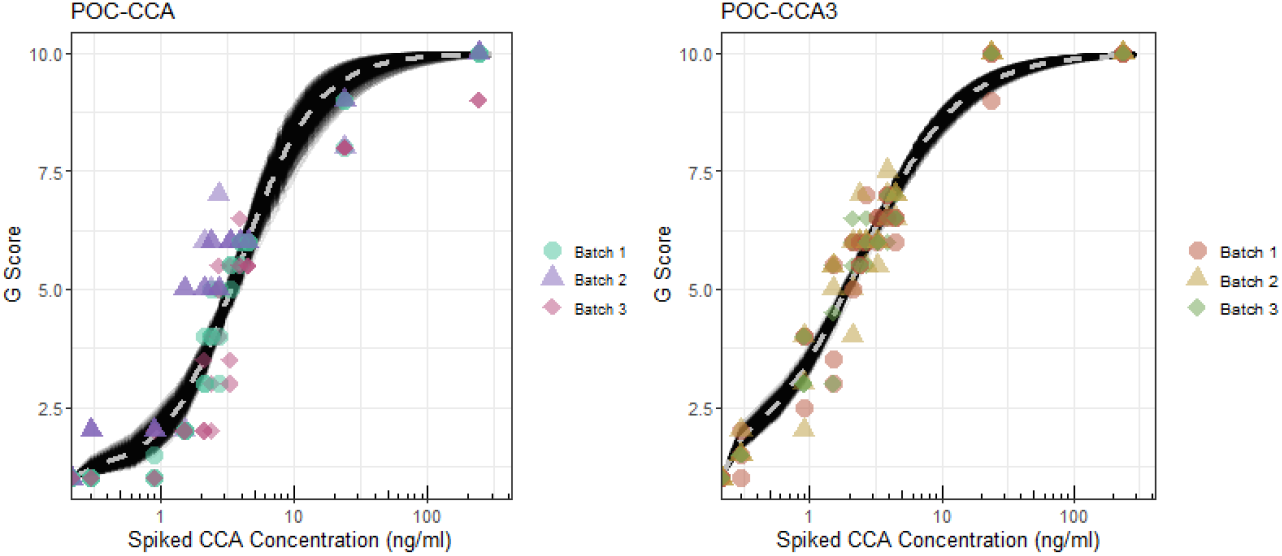
Logistic saturation curves for POC-CCA (left) and POC-CCA3 (right) G-scores for each of the three batches tested against urine spiked with AWA-TCA containing CCA concentrations from 0-240ng/ml. The mean G-score is shown by the dashed grey line.

### 3.2 Field evaluation

Overall, the prevalence was higher in Mayuge than in Tororo, reflecting the endemicity of each setting. KK estimated lower prevalence compared to either POC-CCA or POC-CCA3, irrespective of the G-Score threshold considered using the commonly accepted range of possible thresholds of 2, 3, or 4 (Table 1). The geometric mean of positive egg counts was calculated (not shown) to estimate true prevalence using the pocket chart developed by de Vlas et al. [43]. Using prevalence from all KK slides produces a true prevalence estimate of 30% in Tororo and 95% in Mayuge. Correlation in the average individual-level G-scores between POC-CCA and POC-CCA3 was high in Mayuge (Spearman Rank Correlation 0.75), but much lower in Tororo (Spearman Rank Correlation 0.41). Similarly, the correlation between individual POC-CCA3 G-scores and KK EPG was high in Mayuge (Spearman Rank Correlation 0.66), and lower in Tororo (Spearman Rank Correlation 0.46).

The model predicted a median prevalence of 91.4% in Mayuge and 31.2% in Tororo (Figure 2), considerably higher than estimated by KK in both settings, but comparable to the estimates using average POC-CCA and POC-CCA3, based on a threshold of G3, expect in the case of POC-CCA3 in Mayuge which requires a threshold of G2 (see Table 1).

**Figure 2:**
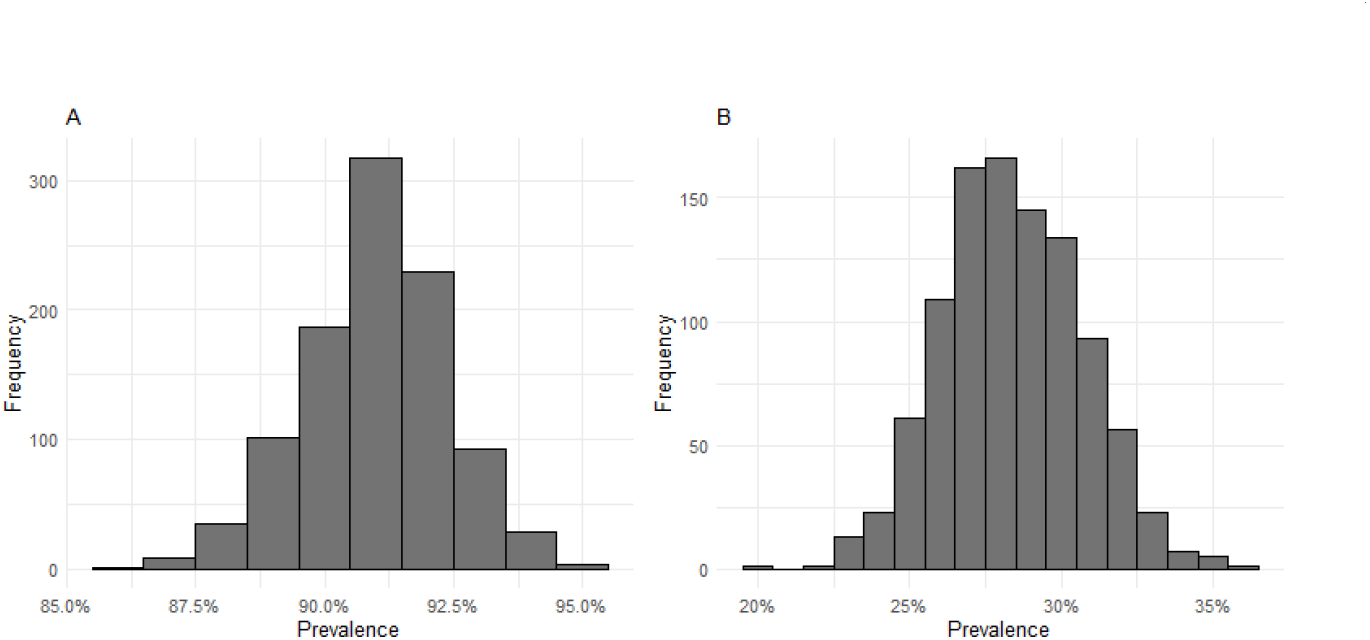
Distribution of model predicted *Schistosoma mansoni* prevalence in Mayuge (A) and Tororo (B).

POC-CCA and POC-CCA3 batch variations using data from the laboratory evaluation (urine samples spiked with CCA) and the field evaluation were distinct in their consistency (Figure 3). Batch-specific variation was estimated to be higher in the field than in the laboratory spiked samples. Overall, POC-CCA3 exhibited minimal batch-specific variation compared to POC-CCA, as indicated by the closer clustering of the variation distributions across batches. The clustering of standard deviation distributions of each batch indicates that G-score differences between batches for the same infection intensity were consistent. POC-CCA3 also demonstrated lower batch-specific variation on average, as indicated by consistently lower batch variation values across all settings. The low standard deviation values indicate limited variability in G-scores produced for the same infection intensity within each batch. Different POC-CCA batches presented high variation in different settings, with batch 3 having notably high batch-specific variation in Mayuge and batch 2 in Tororo, indicating more inconsistent G-scores from that batch.

**Figure 3:**
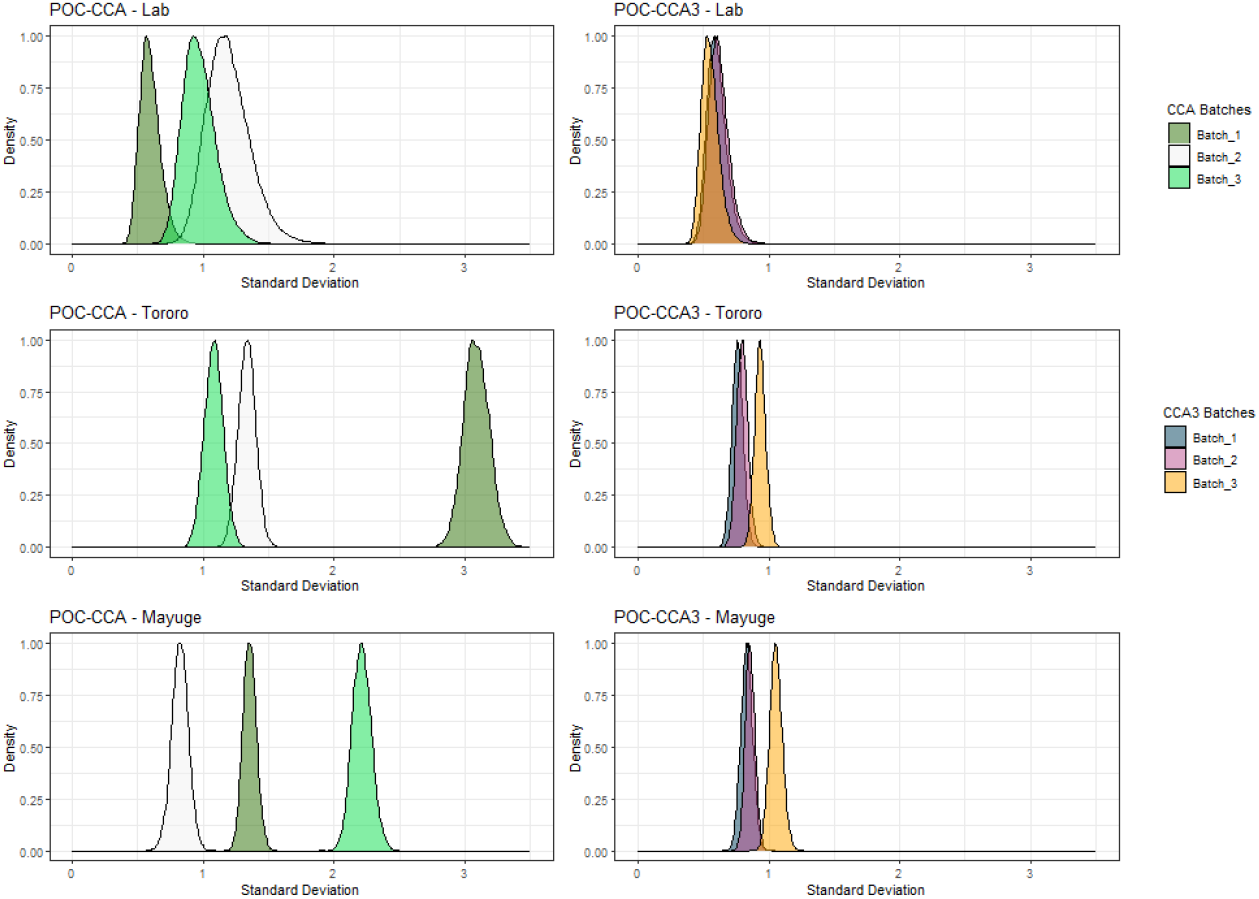
Posterior distributions of the standard deviation (i.e., 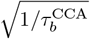 and 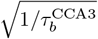) for each batch of POC-CCA (left) and POC-CCA3 (right), estimated under three conditions: laboratory evaluation (using AWA-TCA CCA-spiked urine samples) and field evaluation in Tororo and Mayuge. Each colour denotes a different batch within that setting. A smaller standard deviation indicates less variability in producing G-scores, while larger values suggest greater inconsistency in that batch. The close clustering of the POC-CCA3 batches indicates high batch-to-batch consistency.

POC-CCA3 results from the three pure CCA based calibrators showed increasing G-scores with increasing concentrations, with the differences between the calibrators being statistically significant (F (2, 585) = 106.95, p = 2.6*10-40 OR p < 0.001). However, the difference between G-scores obtained from Calibrator 2 (2.4 ng/mL) and Calibrator 3 (3 ng/mL) was less than 1 G-score unit, Supplemental Material 2. Linear regression (not shown) indicated there was no statistical difference between the POC-CCA3 batches. Comparing the outcomes of the three calibrators across the three POC-CCA3 batches suggests that Calibrator 1 (1.8 ng/mL) can potentially lead to more noise. This occurs around the concentration value where the G-score rapidly increases (see Figure 1). Calibrator 3 led to lower noise across all three

POC-CCA3 batches tested (Supplemental Material 3). Overall, however, each POC-CCA3 batch showed consistent results for each respective calibrator, showing few signs of batch-to-batch variation. The average estimated sensitivity and specificity of POC-CCA3 (1 day sample) with a G-Score threshold of 4 were 86.3% and 92.9%, respectively in Tororo and 88.1% and 95.1% in Mayuge (Figure 4). All values for the estimated sensitivity and specificity values of POC-CCA3 and POC-CCA based on single/multiple sampling and different G-score thresholds can be found in Supplemental Material 4. Sampling more than one day greatly improved the performance of both POC-CCA and POC-CCA3, especially in the lower endemicity setting. POC-CCA3 showed a major advantage over POC-CCA in Tororo, but no significant difference in Mayuge. Compared to KK, POC-CCA and POC-CCA3 outperforms in terms of sensitivity using all thresholds in both settings (Supplemental Material 4 and Supplemental Material 5).

**Figure 4:**
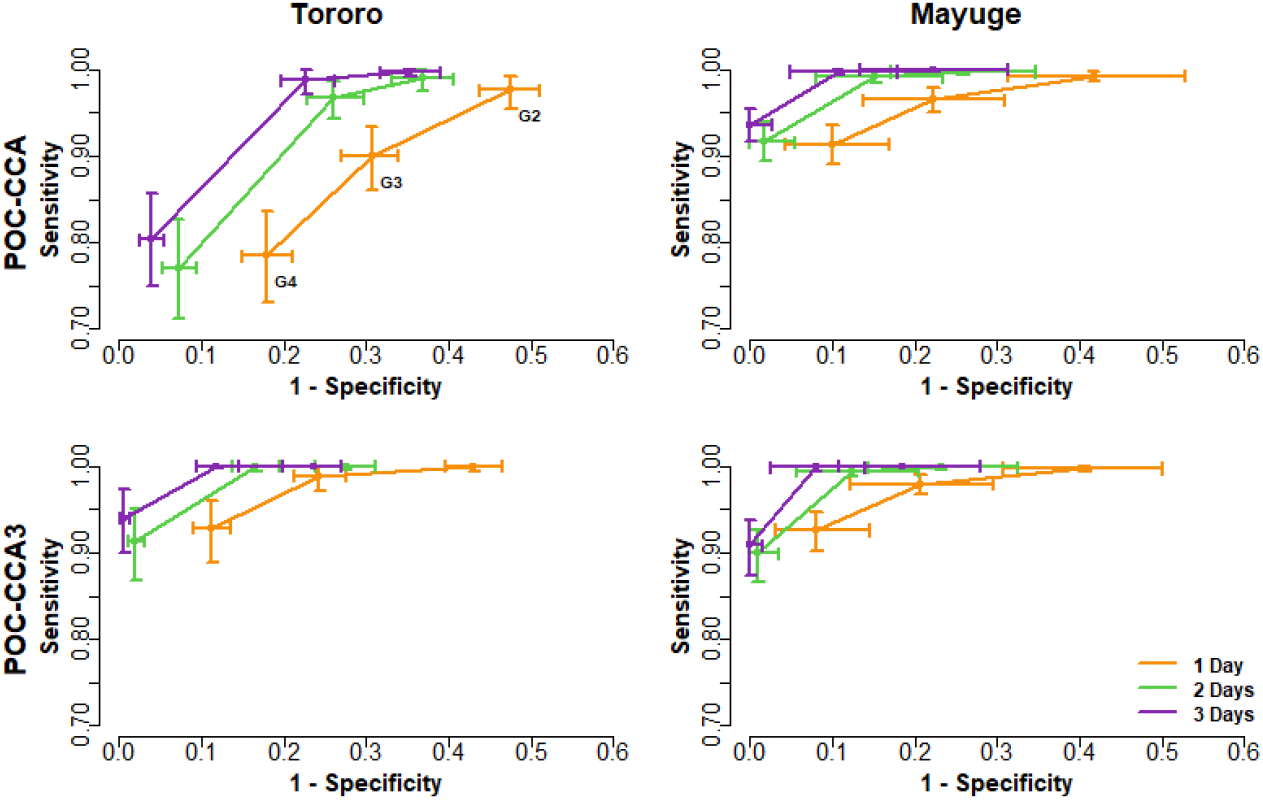
Receiver operating characteristic curves for the POC-CCA (top) and POC-CCA3 (bottom) across the two settings with different Schistosoma mansoni endemicity levels: Tororo (left) and Mayuge (right) have moderate and high endemicity, respectively. Colours represent the number of sampling days (orange—1 day, green—2 days, and purple—3 days). The G-score threshold decreases from left to right (G4, G3, and G2), with one day of sampling only including rounded G-score thresholds.

The AUC, signifying the overall diagnostic performance of the POC-CCA and POC-CCA3 increased for both tests as the number of sampling days increased (Table 2). The POC-CCA3 test had higher median AUC values than the POC-CCA in Tororo, but only slightly higher values were found for POC-CCA3 compared to POC-CCA in Mayuge. Across all settings and sampling days, the increased median AUC values observed with POC-CCA3 were statistically significantly higher than those of POC-CCA, based on a one-tailed, paired Wilcoxon signed-rank test *p <* 2.2 *×* 10^*−*16^.

**Table 2:**
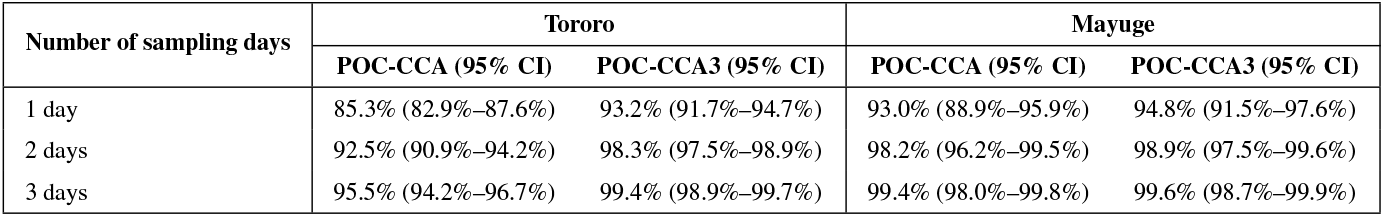
Diagnostic sensitivity of POC-CCA and POC-CCA3 in Tororo and Mayuge across one, two, and three consecutive sampling days. Values represent sensitivity estimates with 95% confidence intervals.

The probability that both POC tests would meet the WHO TPP was expressed as a percentage of simulations (Figure 5). Although the specificity of both tests can be quite high (above 90% depending on the G-score threshold used), the WHO TPP minimum requirement is >95% (for a sample of 100 individuals). A single test in Tororo, with either the original POC-CCA or the POC-CCA3, is unlikely to meet the WHO TPP requirement, irrespective of the G-score threshold used, with both tests failing to achieve TPP. In Mayuge, POC-CCA3 achieved the minimum TPP in 47% of simulations, whereas POC-CCA achieved the minimum TPP in 40% of simulations. Testing two (or better yet, three) days, assuming a G-score threshold of 4, provides a good chance of meeting the minimum TPP requirements (>80% of simulations), Figure 5, with POC-CCA in Tororo as an exception, achieving markedly less even with increased number of samples. POC-CCA displays a markedly reduced probability of achieving the minimum WHO TPP requirements in Tororo compared to POC-CCA3. KK cannot achieve the WHO TPP requirement in a moderate endemic setting, Supplemental Material 5. In a high endemic setting, KK fully achieves the minimum WHO TPP requirement with 1 day of sampling but is unlikely to achieve the optimum WHO TPP requirement even with 3 days of sampling.

**Figure 5:**
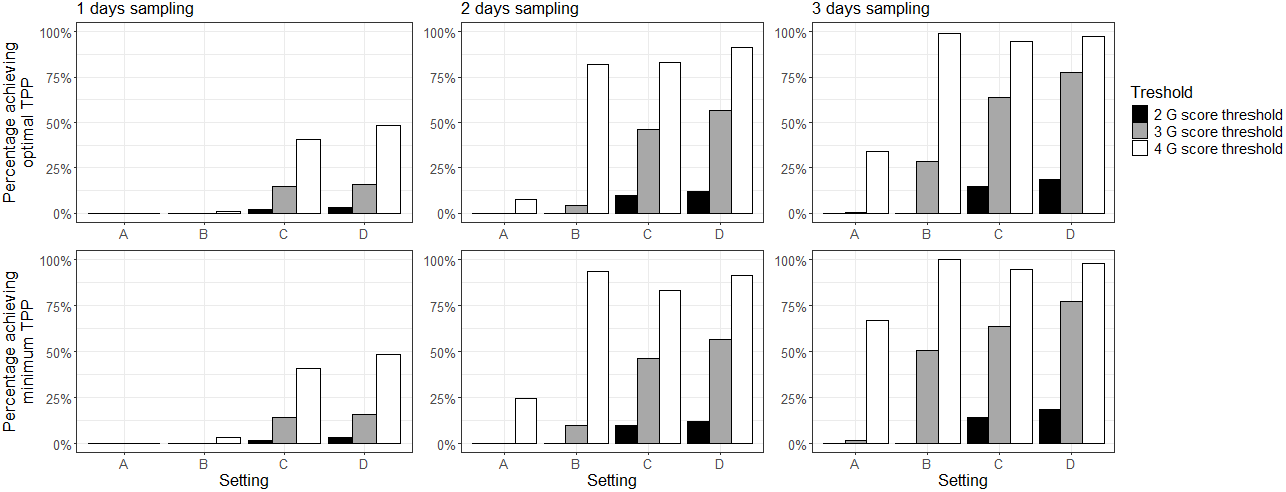
Percentage of simulations where POC-CCA in Tororo (A), POC-CCA3 in Tororo (B), POC-CCA in Mayuge (C), and POC-CCA3 in Mayuge (D), meet the recommended (top) and minimal (bottom) TPP requirement for specificity based on the four thresholds and considering 1, 2 or 3 days of sampling with one test per day.

POC-CCA and POC-CCA3 performed with similar sensitivity and specificity in a setting with 10% prevalence based on KK (using simulations with 20% true prevalence), Table 3. The most accurate G-score threshold predicting the true prevalence was G4.

**Table 3:** Predicted prevalence using POC-CCA or POC-CCA3 in settings with 10% Kato-Katz (KK) prevalence (one slide), with 95% Bayesian Credible Intervals in brackets.

| G-Score | POC-CCA prevalence | POC-CCA3 prevalence | KK prevalence |
| --- | --- | --- | --- |
| 2 | 23.3% (13.0%–52.0%) | 23.0% (12.0%–55.0%) | 10.0% (10.0%–10.0%) |
| 3 | 21.0% (12.0%–38.0%) | 19.5% (10.0%–40.0%) | – |
| 4 | 18.0% (10.0%–28.0%) | 15.8% (8.0%–29.0%) | – |

## 4 Discussion

Batch-to-batch variability issues have been described for the widely used, WHO-endorsed, POC-CCA test [27], prompting the development of an updated version of the test based on recombinant antibodies, combined with enhanced quality control measures in the manufacturing process. Here we evaluated the existing POC-CCA alongside this updated POC-CCA3 in the laboratory and communities endemic for *S. mansoni*. LCA modelling indicates that POC-CCA3 shows minimal batch-to-batch variation, whilst maintaining diagnostic accuracy. Results from the current study indicate that the newly developed POC-CCA3, is therefore an important improvement to the existing widely evaluated and WHO-endorsed POC-CCA. Model predictions indicated that a G-score of 2 provided the most accurate POC-CCA3 prevalence estimates. However, to meet the WHO TPP requirements, a threshold of G-score 4 is recommended, due to the high specificity criteria of the TPP.

### 4.1 Batch-to-batch variation

Data collected in the lab depicting the dose-response relationship highlighted the inconsistent G-scores produced by POC-CCA for the same concentration ranges (Figure 1), confirming ongoing issues with batch-to-batch variation [27]. This is further supported by the posterior distributions produced from the LCA model using laboratory data, which indicate that POC-CCA3 batches exhibit negligible batch-to-batch variation, as demonstrated by the clustering and overlapping of standard deviation distribution (Figure 3), as compared to broader clustering for POC-CCA. Here, batch-specific variation is also shown to be lower in POC-CCA3 demonstrated by the narrower standard deviation distributions depicted by all POC-CCA3 batches.

We evaluated the performance of field calibrators across three batches of POC-CCA3 and found minimal batch-to-batch variation. Although the difference between Calibrators 2 and 3 was very small (<1 average G-score), linear regression analysis indicated no significant difference between batches. While Calibrator 1 (1.8 ng/mL) appeared to introduce greater variability, generally, the consistency observed across POC-CCA3 batches suggests that inclusion of calibrators alongside POC-CCA3 kits is unlikely to be necessary.

Field study data collected in Mayuge and Tororo align with these findings. In both settings POC-CCA3 displays close clustering of batch standard deviation distribution of around 1, whereas POC-CCA displays distinct standard deviation distributions from 1 up to 3, Figure 3. The consistently low standard deviation value of POC-CCA3 is not only indicative of minimal batch-to-batch variation but also the generally reduced batch-specific variation. This means that compared to POC-CCA, the three POC-CCA3 batches generally gave more consistent G-scores values for the same CCA concentrations and intensities of infection, as predicted by the LCA model.

### 4.2 Diagnostic performance

G-scores of both the POC-CCA and POC-CCA3 of all batches increased with known increasing concentrations of AWA-TCA containg CCA-spiked urine samples supporting our understanding of both tests showing semi-quantitative results (Figure 1). Both tests exhibited similar saturation curves while POC-CCA3 demonstrated a more rapid increase at low G-scores compared to the POC-CCA, indicating slightly higher sensitivity at these lower CCA concentrations. This would be an added benefit as more endemic areas move towards elimination and post-MDA surveillance, however for such settings, more sensitive tests are likely needed, and neither POC-CCA nor POC-CCA3 may be sensitive enough [44].

The LCA model predicted a median true prevalence of 91.3% and 31.2% for Mayuge and Tororo respectively, compared with a prevalence of 54.7% and 12.4% respectively based on a single KK slide, aligning with the WHO categories of high (>50% prevalence in school-aged children) and moderate (10-49% prevalence in school-age children) endemicity for these two communities. The model estimates fall in line closely to the true prevalence estimates using the de Vlas pocket chart developed [43]. All diagnostics (single and multiple KK, POC-CCA and POC-CCA3 both at thresholds G2-G4) also classified Mayuge as a highly endemic setting. For Tororo, all diagnostics classified this district as a moderate endemicity setting, except for the POC-CCA when using a threshold of G2 which would have classified Tororo as a high endemicity setting (60.1% prevalence). On a finer scale, however, and consistent with existing literature [45, 46, 47, 48, 49], our findings demonstrate that KK consistently underestimates the prevalence compared to both POC-CCA and POC-CCA3 across G-score thresholds varying between G2 and G4 (Table 1). A comparative analysis of G-scores between POC-CCA and POC-CCA3 revealed a consistently higher average G-score for the latter in both endemic contexts. This is supported by the estimated prevalence between POC-CCA and POC-CCA3 from the first sample, with POC-CCA3 estimating higher prevalences. This, however, changes when considering all samples, where POC-CCA scores indicate higher prevalences, further highlighting the batch-to-batch inconsistency displayed by POC-CCA, with significant differences between estimated prevalences using the first sample only versus a mean of all samples.

The LCA model simulated sensitivity and specificity of a single POC-CCA3 was 88.1% and 95.5%, respectively, in a high endemic setting and 86.3% and 92.9%, respectively, in a moderate endemic setting. The difference between diagnostic performance in moderate and high endemic settings is due to the average infection of intensity of infected individuals in each setting. The ROC analysis and AUC, which summarises the diagnostic accuracy of the different tests, were significantly increased from POC-CCA3 compared to POC-CCA. In the moderate-endemic setting, POC-CCA3 showed a greater improvement in ROC and median AUC (4%–8% difference), while in the high-endemic setting, only a marginal improvement was observed (0.2%–1.8% difference). Ultimately, POC-CCA3 is shown to deliver at least very similar sensitivity and specificity in moderate and endemic settings as POC-CCA.

### 4.3 Target product profile

The WHO has set the TTP threshold at 60% sensitivity and 95% specificity as the minimal requirement and at 75% sensitivity and 96.5% as the recommended requirement for Schistosoma M&E. Optimal G-score thresholds for achieving the WHO TPP was demonstrated to be at G4 for both settings for both POC-CCA and POC-CCA3. A small proportion (<60%) of simulations achieved either the minimal or the recommended TPP with only one day of sampling, even with adjusted G-score thresholds. The main limitation seems to be achieving the >95% specificity in simulations. Due to the very high specificity requirements of the WHO TPP, in most cases, a high G-score threshold is required. However, while achieving the WHO TPP, the true prevalence predicted by the model is underestimated, sacrificing overall accuracy to achieve the required specificity. This is likely due to the POC-CCA and POC-CCA3 tests actually having a higher specificity than even the LCA models predict, as the egg-negative individuals may still inaccurately pull down the denominator, and further work with additional high specificity diagnostics such as egg hatching, CAA testing, and/or stool qPCR may improve the estimates of true infection status and therefore result in improved and potentiallu more accurate specificity measures for the POC-CCA and POC-CCA3 [50]. With three days of sampling, the proportion of simulations achieving the recommended TPP reaches more than 80%, but such a protocol remains logistically and financially unfeasible in M&E settings. Although KK does not face the same specificity challenges as POC-CCA and POC-CCA3, as KK is assumed to have 100% specificity, it can only achieve the minimum WHO TPP requirements in a high-endemic setting due to its reduced sensitivity. (Supplemental Material 5).

### 4.4 Limitations and further works

Some of the diagnostic performance metrics presented, including sensitivity and specificity estimates, are derived from the LCA model, which is informed by laboratory and field datasets. While LCA provides a valuable insight in the absence of a perfect reference standard, it is not without limitations. Ideally, diagnostic performance would be assessed against true gold standards for both sensitivity and specificity. Although KK provides a gold standard for specificity, a true gold standard for sensitivity is currently unavailable for *S. mansoni*, especially in settings of low infection intensity, and hatching and qPCR, as mentioned above would help elucidate this further.

Spiked urine samples were used in the field as calibrators to control for batch-to-batch variation, confirming the small differences as found in the lab. However, the range of CCA concentrations was relatively narrow, resulting in higher noise between Calibrator 1 than from 2 and 3 (see Supplemental Material 3), but this can likely be explained as the calibrator aligns an antigen concentration of 1.8 ng/ml CCA (Supplemental Material 2), which was where the G-score rapidly increased for the POC-CCA3 (see Figure 1). Whilst the G-scores associated with the calibrator sets aligned well with the field G-scores and AWA-TCA and resulting CCA concentrations, for future use it is recommended to extend the concentration range for the calibrators.

Further studies are recommended to evaluate the use of POC-CCA3 with fresh and frozen urine samples, which has previously been demonstrated to affect POC-CCA readings [51, 52]. Additionally, stability studies to better estimate POC-CCA3 shelf life and whether long-term storage after production affects test results would contribute to increasing POC-CCA3 acceptance in the research and disease control community. Cost analyses and production scalability (whilst maintaining improved characteristics) should also be investigated including the ability to potentially produce the improved POC-CCA3 in endemic countries. Given the aims of WHO for EPHP of schistosomiasis by 2030 [5] and the leading role diagnostics will have in guiding intervention decisions, M&E and surveillance, such research should be prioritised.

## 5 Conclusions

The POC-CCA3 test demonstrated minimal batch-to-batch variation and batch-specific variation, greatly improving on the existing POC-CCA. The estimated sensitivity and specificity of a single POC-CCA3 was 88.1% and 95.5%, respectively, in a high endemic setting and 86.3% and 92.9%, respectively, in a moderate endemic setting. To achieve WHO TPP requirements, a G-score threshold of 4 is recommended, however a lower G-score threshold of 2 is required to closely estimate true prevalence. Despite neither the POC-CCA or the POC-CCA3 consistently reach the WHO TPP requirements with a single sample, the POC-CCA remains widely used as one of the best diagnostic alternatives to KK available for *S. mansoni*. POC-CCA3 is a significant improvement that reduces the batch-to-batch variation, whilst maintaining the diagnostic accuracy of the POC-CCA. We therefore strongly endorse its immediate use, alongside further research into test stability, use on frozen samples and work to continue to improve specificity for what is likely the best currently available point-of care diagnostics for *S. mansoni* infections. Market access research and field trials for potentially increasing the use of the POC-CCA3 in other endemic settings and countries are required. This research will produce more data and insights on how the POC-CCA3 can be used to inform control programmes.

## Ethics Statement

Ethics approval for this work was granted by the College Ethics Committee of the College of Medical, Veterinary and Life Sciences, University of Glasgow (Reference: 200220186), and by the Uganda National Council for Science and Technology (UNCST), which approved the research project (Reference: HS2788ES) on 06/07/2023, valid until 20/06/2024. All necessary patient/participant consent (including consent to publish) has been obtained. Identifiers included in the data were not known to anyone outside the research group and cannot be used to identify individuals.

## Data Availability

All scripts used within this study are available online at https://github.com/MabEntez/Schisto-CCA3-LCA. Anonymised data can be shared upon reasonable request from the corresponding author.

## Declarations

All authors confirm that they have the right to post this manuscript and have assented to its submission. All relevant ethical guidelines have been followed. The study involved human subjects and used non-public human data. No prospective interventional study was conducted. All appropriate research reporting guidelines have been followed. The authors are legally responsible for the content of the article.

## References

[1] CDC. About Schistosomiasis, June 2024. URL https://www.cdc.gov/schistosomiasis/about/index.html.

[2] World Health Organization. Schistosomiasis - Fact Sheet, February 2023. URL https://www.who.int/news-room/fact-sheets/detail/schistosomiasis.

[3] World Health Organization. Crossing the billion: Lymphatic filariasis, onchocerciasis, schistosomiasis, soil-transmitted helminthiases and trachoma — preventive chemotherapy for neglected tropical diseases. In Crossing the Billion. 2017. URL https://iris.who.int/handle/10665/255498. Accessed on 27 February 2024.

[4] World Health Organization. Diagnostic target product profiles for monitoring, evaluation and surveillance of schistosomiasis control programmes,. URL https://www.who.int/publications/i/item/9789240031104.

[5] World Health Organization. Ending the neglect to attain the sustainable development goals: A road map for neglected tropical diseases 2021–2030, 2021. URL https://www.who.int/publications-detail-redirect/9789240010352.

[6] Daniel G. Colley, Amaya L. Bustinduy, W. Evan Secor, and Charles H. King. Human schistosomiasis. Lancet (London, England), 383(9936):2253–2264, June 2014. ISSN 1474-547X. doi: 10.1016/S0140-6736(13)61949-2.

[7] Sandra Planchart, Renzo Nino Incani, and Italo Mario Cesari. Preliminary characterization of an adult worm “vomit” preparation of Schistosoma mansoni and its potential use as antigen for diagnosis. Parasitology Research, 101(2):301–309, July 2007. ISSN 0932-0113. doi: 10.1007/s00436-007-0482-2.

[8] Patrick J. Skelly, Akram A. Da’dara, Xiao-Hong Li, William Castro-Borges, and R. Alan Wilson. Schistosome Feeding and Regurgitation. PLOS Pathogens, 10(8):e1004246, August 2014. ISSN 1553-7374. doi: 10.1371/journal.ppat.1004246. URL https://journals.plos.org/plospathogens/article?id=10.1371/journal.ppat.1004246. Publisher: Public Library of Science.

[9] G. J. Van Dam, B. J. Bogitsh, R. J. M. Van Zeyl, J. P. Rotmans, and A. M. Deelder. Schistosoma mansoni: In vitro and In vivo Excretion of CAA and CCA by Developing Schistosomula and Adult Worms. The Journal of Parasitology, 82(4):557, August 1996. ISSN 00223395. doi: 10.2307/3283780. URL https://www.jstor.org/stable/3283780?origin=crossref.

[10] K. S. Warren, E. O. Domingo, and R. B. Cowan. Granuloma formation around schistosome eggs as a manifestation of delayed hypersensitivity. The American Journal of Pathology, 51(5):735–756, November 1967. ISSN 0002-9440. URL https://www.ncbi.nlm.nih.gov/pmc/articles/PMC1965400/.

[11] Joaquín M. Prada, Panayiota Touloupou, Moses Adriko, Edridah M. Tukahebwa, Poppy H. L. Lamberton, and T. Déirdre Hollingsworth. Understanding the relationship between egg- and antigen-based diagnostics of Schistosoma mansoni infection pre- and post-treatment in Uganda. Parasites & Vectors, 11(1):21, January 2018. ISSN 1756-3305. doi: 10.1186/s13071-017-2580-z. URL https://doi.org/10.1186/s13071-017-2580-z.

[12] Suzan C. M. Trienekens, Christina L. Faust, Keila Meginnis, Lucy Pickering, Olivia Ericsson, Andrina Nankasi, Arinaitwe Moses, Edridah M. Tukahebwa, and Poppy H. L. Lamberton. Impacts of host gender on Schistosoma mansoni risk in rural Uganda—A mixed-methods approach. PLOS Neglected Tropical Diseases, 14(5):e0008266, May 2020. ISSN 1935-2735. doi: 10.1371/journal.pntd.0008266. URL https://journals.plos.org/plosntds/article?id=10.1371/journal.pntd.0008266. Publisher: Public Library of Science.

[13] Poppy H. L. Lamberton, Narcis B. Kabatereine, David W. Oguttu, Alan Fenwick, and Joanne P. Webster. Sensitivity and Specificity of Multiple Kato-Katz Thick Smears and a Circulating Cathodic Antigen Test for Schistosoma mansoni Diagnosis Pre- and Post-repeated-Praziquantel Treatment. PLOS Neglected Tropical Diseases, 8(9):e3139, September 2014. ISSN 1935-2735. doi: 10.1371/journal.pntd.0003139. URL https://journals.plos.org/plosntds/article?id=10.1371/journal.pntd.0003139. Publisher: Public Library of Science.

[14] Pytsje T. Hoekstra, Govert J. van Dam, and Lisette van Lieshout. Context-Specific Procedures for the Diagnosis of Human Schistosomiasis – A Mini Review. Frontiers in Tropical Diseases, 2, 2021. ISSN 2673-7515. URL https://www.frontiersin.org/articles/10.3389/fitd.2021.722438.

[15] Paul Ogongo, Thomas M. Kariuki, and R. Alan Wilson. Diagnosis of schistosomiasis mansoni: an evaluation of existing methods and research towards single worm pair detection. Parasitology, 145(11):1355–1366, September 2018. ISSN 0031-1820, 1469-8161. doi: 10.1017/S0031182018000240. URL https://www.cambridge.org/core/journals/parasitology/article/diagnosis-of-schistosomiasis-mansoni-an-evaluation-of-existing-methods-and-research-towards-single-worm-pair-detection/552001BF61F171FB60F11577DC42280A. Number: 11.

[16] Elías Kabbas-Piñango, Moses Arinaitwe, Govert J. van Dam, Adriko Moses, Annet Namukuta, Andrina Barungi Nankasi, Nicholas Khayinja Mwima, Fred Besigye, Joaquin M. Prada, and Poppy H. L. Lamberton. Reproducibility matters: intra- and inter-sample variation of the point-of-care circulating cathodic antigen test in two Schistosoma mansoni endemic areas in Uganda. Philosophical Transactions of the Royal Society B: Biological Sciences, 378(1887):20220275, August 2023. doi: 10.1098/rstb.2022.0275. URL https://royalsocietypublishing.org/doi/10.1098/rstb.2022.0275. Publisher: Royal Society.

[17] Pauline N. M. Mwinzi, Nupur Kittur, Elizabeth Ochola, Philip J. Cooper, Carl H. Campbell, Charles H. King, and Daniel G. Colley. Additional Evaluation of the Point-of-Contact Circulating Cathodic Antigen Assay for Schistosoma mansoni Infection. Frontiers in Public Health, 3, March 2015. ISSN 2296-2565. doi: 10.3389/fpubh.2015.00048. URL https://www.frontiersin.org/journals/public-health/articles/10.3389/fpubh.2015.00048/full. Publisher: Frontiers.

[18] Miriam Casacuberta, Safari Kinunghi, Birgitte J. Vennervald, and Annette Olsen. Evaluation and optimization of the Circulating Cathodic Antigen (POC-CCA) cassette test for detecting Schistosoma mansoni infection by using image analysis in school children in Mwanza Region, Tanzania. Parasite Epidemiology and Control, 1(2):105–115, June 2016. ISSN 2405-6731. doi: 10.1016/j.parepi.2016.04.002. URL https://www.sciencedirect.com/science/article/pii/S2405673115300775.

[19] Louis-Albert Tchuem Tchuenté, Césaire Joris Kueté Fouodo Romuald Isaka Kamwa Ngassam, Laurentine Sumo, Calvine Dongmo Noumedem, Christian Mérimé Kenfack Nestor Feussom Gipwe, Esther Dankoni Nana, J. Russell Stothard, and David Rollinson. Evaluation of Circulating Cathodic Antigen (CCA) Urine-Tests for Diagnosis of Schistosoma mansoni Infection in Cameroon. PLOS Neglected Tropical Diseases, 6(7):e1758, July 2012. ISSN 1935-2735. doi: 10.1371/journal.pntd.0001758. URL https://journals.plos.org/plosntds/article?idw. Publisher: Public Library of Science.

[20] Berhanu Erko, Girmay Medhin, Tilahun Teklehaymanot, Abraham Degarege, and Mengistu Legesse. Evaluation of urine-circulating cathodic antigen (Urine-CCA) cassette test for the detection of Schistosoma mansoni infection in areas of moderate prevalence in Ethiopia. Tropical Medicine & International Health, 18(8):1029–1035, 2013. ISSN 1365-3156. doi: 10.1111/tmi.12117. URL https://onlinelibrary.wiley.com/doi/abs/10.1111/tmi.12117. _eprint: https://onlinelibrary.wiley.com/doi/pdf/10.1111/tmi.12117.

[21] Anthony Danso-Appiah, Jonathan Minton, Daniel Boamah, Joseph Otchere, Richard H. Asmah, Mark Rodgers, Kwabena M. Bosompem, Paolo Eusebi, and Sake J. De Vlas. Accuracy of point-of-care testing for circulatory cathodic antigen in the detection of schistosome infection: systematic review and meta-analysis. BULLETIN OF THE WORLD HEALTH ORGANIZATION, 94(7):522–533, July 2016. ISSN 0042-9686, 1564-0604. doi: 10.2471/BLT.15.158741. URL https://www.webofscience.com/api/gateway?GWVersion=2&SrcAuth=DynamicDOIArticle&SrcApp=WOS&KeyAID=10.2471%2FBLT.15.158741&DestApp=DOI&SrcAppSID=EUW1ED0E4CneTKrXfcy5MZYKI5Ql4&SrcJTitle=BULLETIN+OF+THE+WORLD+HEALTH+ORGANIZATION&DestDOIRegistrantName=WHO+Press. Num Pages: 12 Place: Geneva 27 Publisher: World Health Organization Web of Science ID: WOS:000380733400015.

[22] Hajri Al-Shehri, Artemis Koukounari, Michelle C. Stanton, Moses Adriko, Moses Arinaitwe, Aaron Atuhaire, Narcis B. Kabatereine, and J. Russell Stothard. Surveillance of intestinal schistosomiasis during control: a comparison of four diagnostic tests across five Ugandan primary schools in the Lake Albert region. Parasitology, 145(13):1715–1722, November 2018. ISSN 0031-1820, 1469-8161. doi: 10.1017/S003118201800029X. URL https://www.cambridge.org/core/product/identifier/S003118201800029X/type/journal_article.

[23] Oliver Bärenbold, Amadou Garba, Daniel G. Colley, Fiona M. Fleming, Ayat A. Haggag, Reda M. R. Ramzy, Rufin K. Assaré, Edridah M. Tukahebwa, Jean B. Mbonigaba, Victor Bucumi, Biruck Kebede, Makoy S. Yibi, Aboulaye Meité, Jean T. Coulibaly, Eliézer K. N’Goran, Louis-Albert Tchuem Tchuenté, Pauline Mwinzi, Jürg Utzinger, and Penelope Vounatsou. Translating preventive chemotherapy prevalence thresholds for Schistosoma mansoni from the Kato-Katz technique into the point-of-care circulating cathodic antigen diagnostic test. PLOS Neglected Tropical Diseases, 12(12):e0006941, December 2018. ISSN 1935-2735. doi: 10.1371/journal.pntd.0006941. URL https://dx.plos.org/10.1371/journal.pntd.0006941.

[24] World Health Organization. Eighth report of the Strategic and Technical Advisory Group for Neglected Tropical Diseases (STAG-NTDs),. URL https://www.who.int/publications/m/item/eighth-report-of-the-strategic-and-technical-advisory-group-for-neglected-tropical-diseases-(stag-ntds).

[25] Miriam Casacuberta-Partal, Pytsje T. Hoekstra, Dieuwke Kornelis, Lisette Van Lieshout, and Govert J. Van Dam. An innovative and user-friendly scoring system for standardised quantitative interpretation of the urine-based point-of-care strip test (POC-CCA) for the diagnosis of intestinal schistosomiasis: a proof-of-concept study. Acta Tropica, 199:105150, November 2019. ISSN 0001706X. doi: 10.1016/j.actatropica.2019.105150. URL https://linkinghub.elsevier.com/retrieve/pii/S0001706X19307983.

[26] Jessica Clark, Arinaitwe Moses, Andrina Nankasi, Christina L Faust, Adriko Moses, Diana Ajambo, Fred Besigye, Aaron Atuhaire, Aidah Wamboko, Lauren V Carruthers, Rachel Francoeur, Edridah M Tukahebwa, Joaquin M Prada, and Poppy H L Lamberton. Reconciling Egg- and Antigen-Based Estimates of Schistosoma mansoni Clearance and Reinfection: A Modeling Study. Clinical Infectious Diseases, 74(9):1557–1563, May 2022. ISSN 1058-4838. doi: 10.1093/cid/ciab679. URL https://doi.org/10.1093/cid/ciab679. Number: 9.

[27] Agostinho Gonçalves Viana, Pedro Henrique Gazzinelli-Guimarães, Vanessa Normandio de Castro, Yvanna Louise de Oliveira dos Santos, Ana Cristina Loiola Ruas, Fernando Schemelzer de M. Bezerra, Lilian Lacerda Bueno, Silvio Santana Dolabella, Stefan Michael Geiger, Anna E. Phillips, and Ricardo Toshio Fujiwara. Discrepancy between batches and impact on the sensitivity of point-of-care circulating cathodic antigen tests for Schistosoma mansoni infection. Acta Tropica, 197:105049, September 2019. ISSN 0001-706X. doi: 10.1016/j.actatropica.2019.105049. URL https://www.sciencedirect.com/science/article/pii/S0001706X18313810.

[28] Daniel G. Colley, Reda M.R. Ramzy, Jane Maganga, Safari Kinung’hi, Maurice R. Odiere, Rosemary M. Musuva, and Carl H. Campbell. The POC-CCA assay for detection of Schistosoma mansoni infection needs standardization in production and proper quality control to be reliable. Acta Tropica, 238:106795, February 2023. ISSN 0001706X. doi: 10.1016/j.actatropica.2022.106795. URL https://linkinghub.elsevier.com/retrieve/pii/S0001706X22004867.

[29] Jean T. Coulibaly, Yves K. N’Gbesso, Stefanie Knopp, Nicaise A. N’Guessan, Kigbafori D. Silué, Govert J. Van Dam, Eliézer K. N’Goran, and Jürg Utzinger. Accuracy of Urine Circulating Cathodic Antigen Test for the Diagnosis of Schistosoma mansoni in Preschool-Aged Children before and after Treatment. PLoS Neglected Tropical Diseases, 7(3):e2109, March 2013. ISSN 1935-2735. doi: 10.1371/journal.pntd.0002109. URL https://dx.plos.org/10.1371/journal.pntd.0002109.

[30] Jean T. Coulibaly, Stefanie Knopp, Nicaise A. N’Guessan, Kigbafori D. Silué, Thomas Fürst, Laurent K. Lohourignon, Jean K. Brou, Yve K. N’Gbesso, Penelope Vounatsou, Eliézer K. N’Goran, and Jürg Utzinger. Accuracy of Urine Circulating Cathodic Antigen (CCA) Test for Schistosoma mansoni Diagnosis in Different Settings of Côte d’Ivoire. PLoS Neglected Tropical Diseases, 5(11):e1384, November 2011. ISSN 1935-2735. doi: 10.1371/journal.pntd.0001384. URL https://dx.plos.org/10.1371/journal.pntd.0001384.

[31] Rapid Medical Diagnostics,. URL https://www.rapid-diagnostics.com/.

[32] Govert J. Van Dam, Aldert A. Bergwerff, Jane E. Thomas-Oates, J. Peter Rotmans, Johannis P. Kamerling, Johannes F. G. Vliegenthart, and André M. Deelder. The Immunologically Reactive O-Linked Polysaccharide Chains Derived from Circulating Cathodic Antigen Isolated from the Human Blood Fluke Schistosoma Mansoni have Lewis x as Repeating Unit. European Journal of Biochemistry, 225(1):467–482, October 1994. ISSN 0014-2956, 1432-1033. doi: 10.1111/j.1432-1033.1994.00467.x. URL https://febs.onlinelibrary.wiley.com/doi/10.1111/j.1432-1033.1994.00467.x.

[33] K. Polman, M. M. Diakhate, D. Engels, S. Nahimana, G. J. Van Dam, S.T. Falcão Ferreira, A. M. Deelder, and B. Gryseels. Specificity of circulating antigen detection for schistosomiasis mansoni in Senegal and Burundi. Tropical medicine & international health: TM & IH, 5(8):534–537, August 2000. ISSN 1360-2276. doi: 10.1046/j.1365-3156.2000.00600.x.

[34] Jessica Clark, Arinaitwe Moses, Andrina Nankasi, Christina L. Faust, Moses Adriko, Diana Ajambo, Fred Besigye, Arron Atuhaire, Aidah Wamboko, Candia Rowel, Lauren V. Carruthers, Rachel Francoeur, Edridah M. Tukahebwa, Poppy H. L. Lamberton, and Joaquin M. Prada. Translating From Egg-to Antigen-Based Indicators for Schistosoma mansoni Elimination Targets: A Bayesian Latent Class Analysis Study. Frontiers in Tropical Diseases, 3, 2022. ISSN 2673-7515. URL https://www.frontiersin.org/articles/10.3389/fitd.2022.825721.

[35] S. J. De Vlas, B. Gryseels, G. J. Van Oortmarssen, A. M. Polderman, and J. D. Habbema. A model for variations in single and repeated egg counts in Schistosoma mansoni infections. Parasitology, 104 (Pt 3):451–460, June 1992. ISSN 0031-1820. doi: 10.1017/s003118200006371x.

[36] R Core Team. R: A language and environment for statistical computing., 2020. URL http://www.R-project.org/.

[37] JAGS: A program for analysis of Bayesian graphical models using Gibbs sampling. – ScienceOpen,. URL https://www.scienceopen.com/document?vid=ea9f34c2-6244-46d9-b128-831c105999aa.

[38] Matthew J. Denwood. runjags: An R Package Providing Interface Utilities, Model Templates, Parallel Computing Methods and Additional Distributions for MCMC Models in JAGS. Journal of Statistical Software, 71:1–25, July 2016. ISSN 1548-7660. doi: 10.18637/jss.v071.i09. URL https://doi.org/10.18637/jss.v071.i09.

[39] Lisette Van Etten, Dirk Engels, Frederik W. Krijger, Leopold Nkulikyinka, Bruno Gryseels, and Andre M. Deelder. Fluctuation of Schistosome Circulating antigen Levels in Urine of Individuals with Schistosoma mansoni Infection in Burundi. April 1996. doi: 10.4269/ajtmh.1996.54.348. URL https://www.ajtmh.org/view/journals/tpmd/54/4/article-p348.xml.

[40] Govert J. Van Dam, Peter Odermatt, Luz Acosta, Robert Bergquist, Claudia J. De Dood, Dieuwke Kornelis, Sinuon Muth, Jürg Utzinger, and Paul L.A.M. Corstjens. Evaluation of banked urine samples for the detection of circulating anodic and cathodic antigens in Schistosoma mekongi and S. japonicum infections: A proof-of-concept study. Acta Tropica, 141:198–203, January 2015. ISSN 0001706X. doi: 10.1016/j.actatropica.2014.09.003. URL https://linkinghub.elsevier.com/retrieve/pii/S0001706X14002940.

[41] N. Midzi, A. E. Butterworth, T. Mduluza, S. Munyati, A. M. Deelder, and G. J. van Dam. Use of circulating cathodic antigen strips for the diagnosis of urinary schistosomiasis. Transactions of the Royal Society of Tropical Medicine and Hygiene, 103(1):45–51, January 2009. ISSN 1878-3503. doi: 10.1016/j.trstmh.2008.08.018.

[42] D. De Clercq, M. Sacko, J. Vercruysse, V. vanden Bussche, A. Landouré, A. Diarra, B. Gryseels, and A. Deelder. Circulating anodic and cathodic antigen in serum and urine of mixed Schistosoma haematobium and S. mansoni infections in Office du Niger, Mali. Tropical medicine & international health: TM & IH, 2(7):680–685, July 1997. ISSN 1360-2276. doi: 10.1046/j.1365-3156.1997.d01-354.x.

[43] S. J. de Vlas, B. Gryseels, G. J. van Oortmarssen, A. M. Polderman, and J. D. F. Habbema. A pocket chart to estimate true Schistosoma mansoni prevalences. Parasitology Today, 9(8):306–307, August 1993. ISSN 0169-4758. doi: 10.1016/0169-4758(93)90132-Y. URL https://www.sciencedirect.com/science/article/pii/016947589390132Y.

[44] Mariana Silva Sousa, Govert J. Van Dam, Marta Cristhiany Cunha Pinheiro, Claudia J. De Dood, Jose Mauro Peralta, Regina Helena Saramago Peralta, Elizabeth De Francesco Daher, Paul L. A. M. Corstjens, and Fernando Schemelzer Moraes Bezerra. Performance of an Ultra-Sensitive Assay Targeting the Circulating Anodic Antigen (CAA) for Detection of Schistosoma mansoni Infection in a Low Endemic Area in Brazil. Frontiers in Immunology, 10:682, April 2019. ISSN 1664-3224. doi: 10.3389/fimmu.2019.00682. URL https://www.frontiersin.org/article/10.3389/fimmu.2019.00682/full.

[45] Edwige A. Sokouri, Bernardin Ahouty, Innocent A. Abé, Flora G. D. Yao, Thomas K. Konan, Oscar A. Nyangiri, Annette MacLeod, Enock Matovu, Harry Noyes, and Mathurin Koffi. Evaluation of the epidemiological situation of intestinal schistosomiasis using the POC-CCA parasite antigen test and the Kato-Katz egg count test in school-age children in endemic villages in western Côte d’Ivoire. Parasite, 31:66, 2024. ISSN 1776-1042. doi: 10.1051/parasite/2024049. URL https://www.parasite-journal.org/articles/parasite/abs/2024/01/parasite230147/parasite230147.html.

[46] Catieli Gobetti Lindholz, Vivian Favero, Carolina de Marco Verissimo, Renata Russo Frasca Candido, Renata Perotto de Souza, Renata Rosa dos Santos, Alessandra Loureiro Morassutti, Helio Radke Bittencourt, Malcolm K. Jones, Timothy G. St Pierre, and Carlos Graeff-Teixeira. Study of diagnostic accuracy of Helmintex, Kato-Katz, and POC-CCA methods for diagnosing intestinal schistosomiasis in Candeal, a low intensity transmission area in northeastern Brazil. PLOS Neglected Tropical Diseases, 12(3):e0006274, March 2018. ISSN 1935-2735. doi: 10.1371/journal.pntd.0006274. URL https://journals.plos.org/plosntds/article?id=10.1371/journal.pntd.0006274.

[47] Benard Chieng, Collins Okoyo, Elses Simiyu, Paul Gichuki, Cassian Mwatele, Stella Kepha, Sammy Njenga, and David Mburu. Comparison of quantitative polymerase chain reaction, Kato-Katz and circulating cathodic antigen rapid test for the diagnosis of Schistosoma mansoni infection: A cross-sectional study in Kirinyaga County, Kenya. Current Research in Parasitology & Vector-Borne Diseases, 1:100029, January 2021. ISSN 2667-114X. doi: 10.1016/j.crpvbd.2021.100029. URL https://www.sciencedirect.com/science/article/pii/S2667114X21000236.

[48] Nupur Kittur, Jennifer D. Castleman, Carl H. Campbell, Charles H. King, and Daniel G. Colley. Comparison of Schistosoma mansoni Prevalence and Intensity of Infection, as Determined by the Circulating Cathodic Antigen Urine Assay or by the Kato-Katz Fecal Assay: A Systematic Review. March 2016. doi: 10.4269/ajtmh.15-0725. URL https://www.ajtmh.org/view/journals/tpmd/94/3/article-p605.xml.

[49] Warllem Junio Oliveira, Fernanda do Carmo Magalhães, Andressa Mariana Saldanha Elias, Vanessa Normandio de Castro, Vivian Favero, Catieli Gobetti Lindholz, Áureo Almeida Oliveira, Fernando Sergio Barbosa, Frederico Gil, Maria Aparecida Gomes, Carlos Graeff-Teixeira, Martin Johannes Enk, Paulo Marcos Zech Coelho, Mariângela Carneiro, Deborah Aparecida Negrão-Corrêa, and Stefan Michael Geiger. Evaluation of diagnostic methods for the detection of intestinal schistosomiasis in endemic areas with low parasite loads: Saline gradient, Helmintex, Kato-Katz and rapid urine test. PLOS Neglected Tropical Diseases, 12(2):e0006232, February 2018. ISSN 1935-2735. doi: 10.1371/journal.pntd.0006232. URL https://journals.plos.org/plosntds/article?id=10.1371/journal.pntd.0006232.

[50] J. Utzinger, S. L. Becker, L. van Lieshout, G. J. van Dam, and S. Knopp. New diagnostic tools in schistosomiasis. Clinical Microbiology and Infection, 21(6):529–542, June 2015. ISSN 1198-743X. doi: 10.1016/j.cmi.2015.03.014. URL https://www.sciencedirect.com/science/article/pii/S1198743X1500378X.

[51] Estelle Mezajou Mewamba, Arnol Auvaker Zebaze Tiofack, Cyrille Nguemnang Kamdem, Romuald Isaka Kamwa Ngassam, Mureille Carole Tchami Mbagnia, Oscar Nyangiri, Harry Noyes, Hilaire Marcaire Womeni, Flobert Njiokou, and Gustave Simo. Field assessment in Cameroon of a reader of POC-CCA lateral flow strips for the quantification of Schistosoma mansoni circulating cathodic antigen in urine. PLOS Neglected Tropical Diseases, 15(7):e0009569, July 2021. ISSN 1935-2735. doi: 10.1371/journal.pntd.0009569. URL https://dx.plos.org/10.1371/journal.pntd.0009569.

[52] Tereza Cristina Favre, Lilian Christina Nóbrega Holsback Beck, Fernando Schemelzer Moraes Bezerra, Carlos Graeff-Teixeira, Paulo Marcos Zech Coelho, Martin Johannes Enk, Naftale Katz, Ricardo Riccio Oliveira, Mitermayer Galvão Dos Reis, and Otávio Sarmento Pieri. Reliability of point-of-care circulating cathodic antigen assay for diagnosing schistosomiasis mansoni in urine samples from an endemic area of Brazil after one year of storage at −20 degrees Celsius. Revista da Sociedade Brasileira de Medicina Tropical, 55:e0389–2021, 2022. ISSN 1678-9849, 0037-8682. doi: 10.1590/0037-8682-0389-2021. URL http://www.scielo.br/scielo.php?script=sci_arttext&pid=S0037-86822022000100303&tlng=en.

