## Supplementary Materials for "POC-CCA3: Reducing batch-to-batch variation in the WHO-endorsed POC-CCA for *Schistosoma mansoni* and improving test interpretation"

**Supplementary:**
Use of calibrators for the POC-CCA3 in the field

**Supplemental Material 1.** Allocation of urine samples to diagnostic test batches across three consecutive sampling days for each patient group (A–C). On Day 1, each sample was tested with all three batches of both the POC-CCA and POC-CCA3 tests. On Days 2 and 3, a single batch—randomly assigned per sample—was used consistently for each test. In cases where a specific batch was unavailable, only two batches were used. This variability was accounted for in the analytical model. For the KK diagnostic, each stool sample was tested in duplicate (Replicates 1 and 2) on each sampling day.

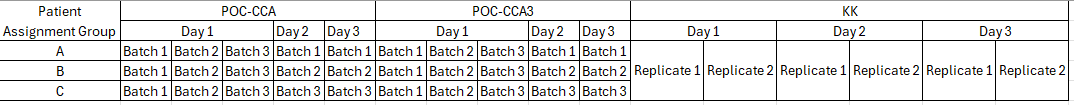

**Supplemental Material 2.** Spiked CCA concentrations, associated adult worm concentrations and G-scores for all field tested POC-CCA3., SE: Standard error

|  | **CCA (ng/mL)** | **G-score (mean ± SE)** |
| --- | --- | --- |
| Calibrator 1 | 1.8 | 4.4 ± 0.1 |
| Calibrator 2 | 2.4 | 6.1 ± 0.1 |
| Calibrator 3 | 3 | 6.5 ± 0.1 |

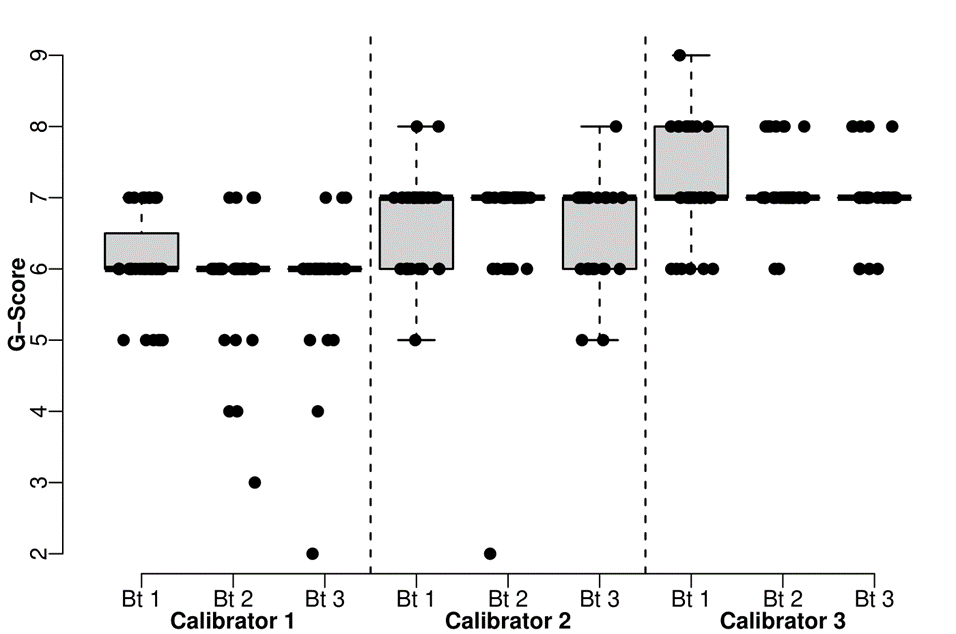

**Supplemental Material 3.** Boxplot of the G-scores of POC-CCA3 across the three Calibrators (1-3) and the three batches (Bt 1-3).

| **Setting** | **Diagnostic** | **Days of sampling** | **G-score Threshold** | **Sensitivity (95% CI)** | **Specificity (95% CI)** |
| --- | --- | --- | --- | --- | --- |
| Tororo | POC-CCA3 | 1 Day | 2 | 100.0% (99.3% - 100.0%) | 57.0% (53.4% - 60.4%) |
|  |  |  | 3 | 98.9% (97.2% - 100.0%) | 75.8% (72.4% - 78.8%) |
|  |  |  | 4 | 92.8% (88.9% - 96.0%) | 88.7% (86.4% - 91.0%) |
|  |  | 2 Days | 2 | 100.0% (99.7% - 100.0%) | 72.5% (68.9% - 76.2%) |
|  |  |  | 3 | 99.3% (98.1% - 100.0%) | 90.9% (88.8% - 93.0%) |
|  |  |  | 4 | 91.3% (86.9% - 95.1%) | 98.1% (97.0% - 99.0%) |
|  |  | 3 Days | 2 | 100.0% (100.0% - 100.0%) | 76.4% (73.0% - 80.1%) |
|  |  |  | 3 | 100.0% (99.2% - 100.0%) | 94.8% (93.0% - 96.4%) |
|  |  |  | 4 | 94.0% (90.0% - 97.3%) | 99.4% (98.8% - 99.9%) |
|  | POC-CCA | 1 Day | 2 | 97.7% (95.4% - 99.2%) | 52.5% (48.9% - 56.2%) |
|  |  |  | 3 | 90.1% (86.1% - 93.4%) | 69.3% (66.0% - 73.1%) |
|  |  |  | 4 | 78.5% (73.1% - 83.5%) | 82.1% (79.0% - 85.1%) |
|  |  | 2 Days | 2 | 99.0% (97.6% - 100.0%) | 63.2% (59.5% - 66.9%) |
|  |  |  | 3 | 92.4% (88.7% - 95.6%) | 82.4% (79.1% - 85.2%) |
|  |  |  | 4 | 77.1% (71.2% - 82.7%) | 92.9% (90.7% - 94.7%) |
|  |  | 3 Days | 2 | 99.7% (99.2% - 100.0%) | 64.8% (61.1% - 68.3%) |
|  |  |  | 3 | 95.8% (92.9% - 97.9%) | 86.4% (83.6% - 89.1%) |
|  |  |  | 4 | 80.3% (75.0% - 85.7%) | 96.0% (94.5% - 97.4%) |
| Mayuge | POC-CCA3 | 1 Day | 2 | 99.8% (99.3% - 100.0%) | 59.4% (50.0% - 69.2%) |
|  |  |  | 3 | 98.0% (96.8% - 99.0%) | 79.4% (70.4% - 87.8%) |
|  |  |  | 4 | 92.6% (90.1% - 94.7%) | 92.0% (85.4% - 96.9%) |
|  |  | 2 Days | 2 | 99.9% (99.7% - 100.0%) | 76.8% (67.4% - 85.7%) |
|  |  |  | 3 | 98.1% (96.7% - 99.1%) | 94.3% (88.3% - 98.8%) |
|  |  |  | 4 | 89.9% (86.6% - 92.7%) | 99.1% (96.6% - 100.0%) |
|  |  | 3 Days | 2 | 100.0% (99.9% - 100.0%) | 81.5% (72.2% - 89.3%) |
|  |  |  | 3 | 99.0% (97.9% - 99.7%) | 97.5% (93.0% - 100.0%) |
|  |  |  | 4 | 90.8% (87.3% - 93.7%) | 100.0% (98.6% - 100.0%) |
|  | POC-CCA | 1 Day | 2 | 99.2% (98.7% - 99.8%) | 58.3% (47.3% - 68.8%) |
|  |  |  | 3 | 96.5% (95.1% - 97.8%) | 77.8% (69.1% - 86.3%) |
|  |  |  | 4 | 91.3% (89.1% - 93.5%) | 90.0% (83.0% - 95.7%) |
|  |  | 2 Days | 2 | 99.8% (99.4% - 100.0%) | 74.2% (65.4% - 83.0%) |
|  |  |  | 3 | 97.8% (96.7% - 98.8%) | 91.9% (85.5% - 97.3%) |
|  |  |  | 4 | 91.6% (89.4% - 93.9%) | 98.3% (94.6% - 100.0%) |
|  |  | 3 Days | 2 | 100.0% (99.8% - 100.0%) | 77.8% (68.6% - 86.7%) |
|  |  |  | 3 | 99.0% (98.2% - 99.6%) | 95.5% (90.6% - 99.0%) |
|  |  |  | 4 | 93.6% (91.6% - 95.4%) | 100.0% (97.4% - 100.0%) |

**Supplemental Material 4**. Simulated sensitivity and specificity estimates expressed as mean (95% credible intervals) for POC‐CCA3 and POC‐CCA diagnostics at G-Score thresholds of 2, 3 and 4, based on model posteriors from Mayuge and Tororo.

| Setting | Days of sampling | KK Sensitivity |
| --- | --- | --- |
| Tororo | 1 Day | 46.7% (38.9% - 54.6%) |
| Tororo | 2 Days | 45.7% (37.9% - 54.1%) |
| Tororo | 3 Days | 45.3% (37.5% - 53.9%) |
| Mayuge | 1 Day | 66.8% (62.4% - 71.3%) |
| Mayuge | 2 Days | 71.1% (67.0% - 75.4%) |
| Mayuge | 3 Days | 72.6% (68.3% - 76.9%) |

**Supplemental Material 5.** Simulated Kato-Katz sensitivity using model posteriors from both Mayuge and Tororo with 95% Credible Intervals in brackets. Kato-Katz specificity is assumed to be 100%.

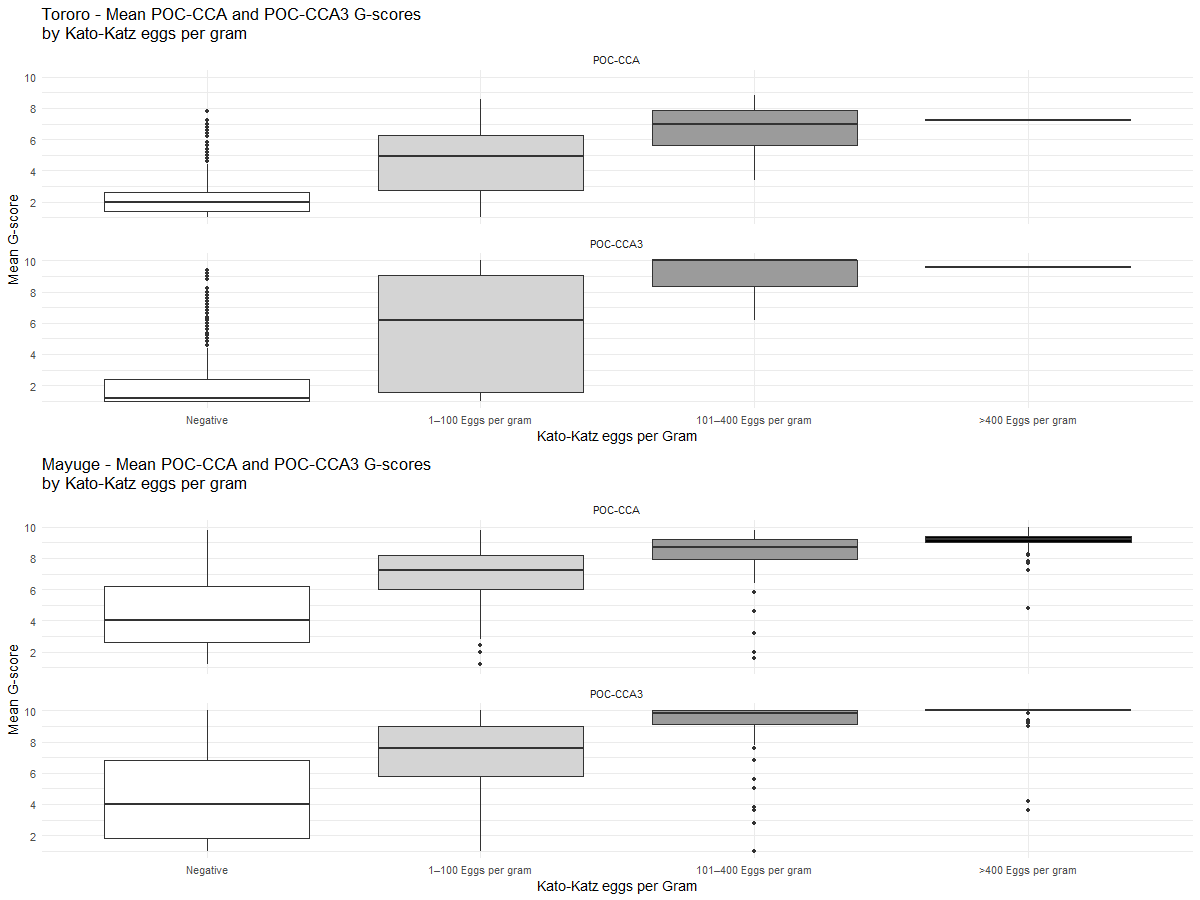

**Supplementary Materials 6.** Boxplots of POC-CCA and POC-CCA3 G-score distributions stratified by Kato–Katz infection intensity category (negative; 1–100; 101–400; >400 eggs per gram. Top row shows Tororo data, bottom row Mayuge data.
